# Barriers to Vaccine Uptake Among Older Adults: A Qualitative Community-Engaged Study

**DOI:** 10.64898/2026.01.13.26344048

**Authors:** Virginie Zoumenou, Verona Mulgrave, Dionne Ray, Kritika Gupta

## Abstract

**Background:** Vaccine-preventable diseases pose significant health risks to older adults. Despite widespread vaccine availability, hesitancy related to uncertainty, misinformation, and access challenges continues to affect vaccination uptake, presenting an ongoing public health concern.

**Objective:** This study aimed to assess perceptions related to vaccination, develop age-appropriate educational strategies through community engagement, and evaluate changes in vaccine-related attitudes following an educational intervention among older adults in rural Delaware and Maryland.

**Study Design:** A mixed-methods approach was employed, combining quantitative surveys with qualitative focus groups and interviews to examine vaccination-related perceptions and experiences.

**Participants:** The study included 124 participants prior to the intervention and 89 participants following the intervention. Participants were older adults residing in Sussex County (Delaware) and Somerset and Wicomico Counties (Maryland).

**Analysis:** Quantitative data were analyzed using logistic regression, while qualitative data were analyzed thematically using inductive coding techniques. Comparisons between pre-and post-intervention findings examined changes in reported barriers, facilitators, perceptions, and confidence related to vaccination.

**Results:** Prior to the intervention, limited access to clear and reliable information (57.4%) and uncertainty toward public health guidance (36.8%) were commonly reported barriers, while family support and healthcare provider recommendations were identified as key facilitators. Following the intervention, reported barriers to vaccination decreased by 93%, and confidence in healthcare provider guidance increased by 190%. Educational materials were well received, with participants reporting improved understanding of vaccine effectiveness and safety, although some hesitation remained.

**Conclusion:** Educational interventions delivered through community-engaged approaches were associated with reduced barriers and increased confidence in vaccination among older adults. These findings highlight the value of targeted health education in supporting informed decision-making and suggest the need for continued public health efforts to sustain vaccine confidence. Further research is warranted to assess long-term outcomes and applicability across additional settings.

## BACKGROUND

Vaccine-preventable diseases continue to pose a substantial global health burden, particularly among older adults, where they are associated with adverse outcomes such as absenteeism, lost productivity, functional decline, disability, and mortality (Ecarnot et al., 2020; Sundaram et al., 2015). A key challenge in preventing these diseases is vaccine hesitancy, defined as the delay in acceptance or refusal of vaccination despite availability (Dubé et al., 2013). Vaccine hesitancy complicates disease prevention efforts and is influenced by a range of factors, including religious beliefs, cultural norms, complacency, convenience, confidence in vaccines, and individuals’ assessment of perceived benefits and risks (Sundaram et al., 2015). Among children and young adults, parental or guardian hesitancy or refusal further contributes to suboptimal vaccination coverage (Crosby et al., 2023; Santibanez et al., 2020).

The determinants of vaccination behavior have been conceptualized using the 5As taxonomy, which identifies access, affordability, awareness, acceptance, and activation as key domains influencing uptake (Thomson et al., 2016). Research focused on older adults highlights multiple barriers to vaccination, including uncertainty toward healthcare systems, misinformation, and perceptions of limited vaccine effectiveness (Bhanu et al., 2021).

Additional factors such as socio-demographic characteristics, logistical constraints, prevailing attitudes toward prevention, and vaccine hesitancy—shaped by complacency, convenience, confidence, and emotional responses amplified through social media—further affect vaccination decisions (Ecarnot et al., 2020). Safety concerns, financial costs, lack of insurance coverage, and absence of healthcare provider recommendations have also been identified as significant barriers among older adults (Kolobova et al., 2022; Moosa et al., 2022).

Conversely, several facilitators have been associated with higher vaccination uptake. These include recommendations from trusted healthcare professionals, supportive social networks, increasing age, presence of chronic conditions, engagement in preventive health behaviors, regular healthcare utilization, and prior vaccination history (Bhanu et al., 2021; Moosa et al., 2022; Roller-Wirnsberger et al., 2021; Yeung et al., 2016). Contextual factors such as urban residence, health literacy, economic stability, and community characteristics have also been linked to increased pneumococcal vaccination rates (Gatwood et al., 2020). Importantly, perceived vaccine efficacy and safety have been shown to influence vaccination decisions more strongly than knowledge alone (Yeung et al., 2016). Previous studies emphasize the importance of comprehensive education efforts, increased public awareness, engagement of healthcare professionals, and the reduction of financial and structural barriers through policy and programmatic support (Kolobova et al., 2022). Advice from healthcare providers, family members, and peers, as well as no-cost vaccination opportunities, further facilitate uptake (Yeung et al., 2016). Targeted outreach delivered through healthcare workers has been effective in addressing hesitancy and increasing COVID-19 vaccination among older adults in community settings, as demonstrated in Singapore (Moosa et al., 2022).

Historically, limited engagement in preventive care, including vaccination, has been observed even when services are readily available. This pattern may reflect limited understanding of preventive effectiveness, a tendency to discount future health benefits, or concerns related to safety and potential consequences. Cultural beliefs and confidence in health systems may also shape vaccination decisions, including apprehension about adverse effects or perceived social implications of vaccination. Awareness interventions have therefore been recognized as a critical component of successful vaccination strategies. Public awareness plays an essential role in encouraging older adults to engage in preventive behaviors and supporting communities in assuming greater responsibility for health protection. However, such interventions must be strategically designed and adequately resourced to communicate clear, consistent messages and demonstrate the practical feasibility of vaccination.

Understanding the factors that influence vaccination decisions among older adults living in rural settings is essential for developing effective, context-appropriate strategies to improve vaccine uptake. This study was designed to examine these factors and to compare vaccination-related perceptions and behaviors among older adults residing in town and city settings in rural areas of Delaware and Maryland.

Study Objectives

The study had three primary objectives:

1. To assess participants’ perceptions related to vaccination.
2. To develop age-appropriate educational intervention strategies, informed by community engagement, to increase confidence in vaccination and reduce hesitancy.
3. To evaluate changes in participants’ vaccination-related perceptions following the awareness intervention.

## METHODS

### Study design, research context, and population

#### Study design

This study used a mixed-methods approach, combining quantitative and qualitative research methods. For the quantitative data, a survey was conducted to capture demographics, vaccination status of children and adults, and barriers to vaccine uptake. For the qualitative data, focus groups (n=5) and interviews (n=29) were conducted to capture deeper insights into barriers and facilitators to vaccine uptake in older adults.

#### Population

The selected rural counties for this study, Sussex County in Delaware and Somerset and Wicomico counties in Maryland, were identified based on several criteria, including County health ranking, Pandemic Vulnerability Index (PVI), vaccination rates, Social Vulnerability Index (SVI), demographic, and community classification as town or city (Table 1). Sussex County in Southern Delaware State shares borders with Wicomico County in the Lower Eastern Shore (LES) of Maryland. Wicomico County shares borders with Somerset County in LES. Counties were also selected because of the existing trust built through Extension programs, including the Parenting Program in Sussex County, Delaware; the Nutrition and Health Programs; and the Well-Connected Communities (WCC)– Culture of Health Initiative on the Lower Eastern Shore of Maryland.

**Table 1.**
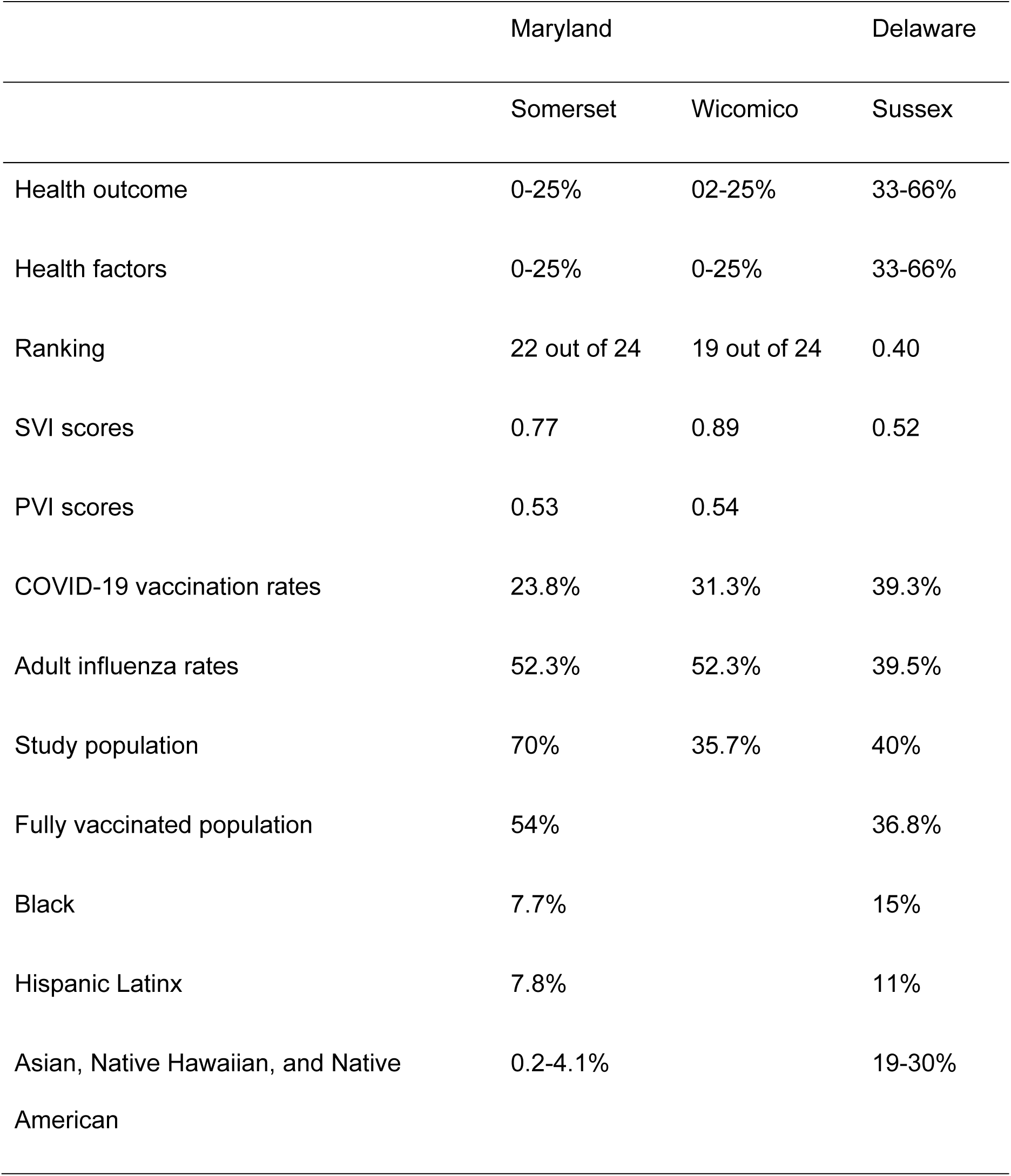
Health factors and vaccination rates in Maryland (Somerset and Wicomo counties) and Delaware (Sussex County)

|  | Maryland |  | Delaware |
| --- | --- | --- | --- |
|  | Somerset | Wicomico | Sussex |
| Health outcome | 0-25% | 02-25% | 33-66% |
| Health factors | 0-25% | 0-25% | 33-66% |
| Ranking | 22 out of 24 | 19 out of 24 | 0.40 |
| SVI scores | 0.77 | 0.89 | 0.52 |
| PVI scores | 0.53 | 0.54 |  |
| COVID-19 vaccination rates | 23.8% | 31.3% | 39.3% |
| Adult influenza rates | 52.3% | 52.3% | 39.5% |
| Study population | 70% | 35.7% | 40% |
| Fully vaccinated population | 54% |  | 36.8% |
| Black | 7.7% |  | 15% |
| Hispanic Latinx | 7.8% |  | 11% |
| Asian, Native Hawaiian, and Native American | 0.2-4.1% |  | 19-30% |

### Data collection

This study was conducted in three phases. Phase I involved coordination among existing coalitions, particularly the coalition built by the UMES Well-Connected Community and all the organizations working with FCS and 4-H in Sussex County, Delaware. Youth and adults from each community were trained to use the Rapid Community Assessment Tool, based on the WCC experience. Post-training, the community members conducted their own assessments and shared the results with the communities. Upon receiving the results, the youth and adults in the community, in collaboration with the Health Council, brainstormed strategies that could work in their community. The research team supported all training, listening sessions, and interview facilitation. To increase participation and compensate participants for their time, incentives were provided. Data analysis details are provided later in this section.

Phase II involved strategy implementation. Each strategy was age- and culturally appropriate (18-29; 30-59; 60-65). The educational tools were made available in both Spanish and English. Dissemination was implemented through social services, Extension program activities, social media channels, churches, community-based organizations, etc. Figure 1 presents the poster used as an educational tool. Phase III provided an opportunity to conduct a rapid assessment after Phase II strategy implementation. The major objective of Phase III was to assess improvement in participant attitudes and behaviors towards vaccination.

**Figure 1.**
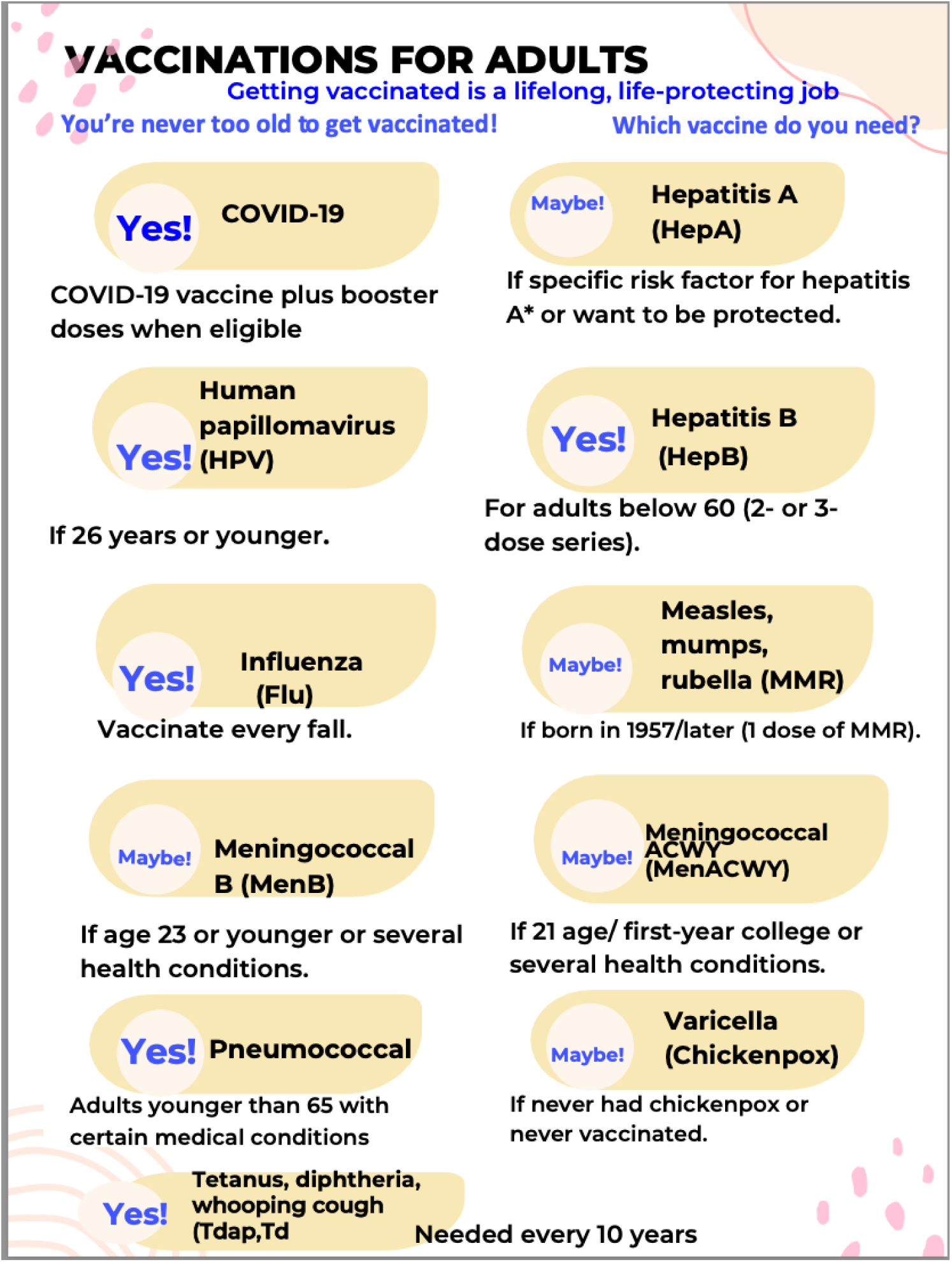
Educational tool used in this study (Adapted from Vaccinations for Adults You’re never too old to get vaccinated! Immunize.org) (Vaccinations for Adults, n.d.)

### Study tools

#### Discussion guide for interviews and focus groups

A semi-structured discussion guide was developed collaboratively by the research team to explore students’ perceptions of health, well-being, and campus resource utilization. The initial draft was prepared by three researchers with advanced training in qualitative methods, public health research, and program evaluation, including two doctoral-level investigators and one graduate-level researcher.

Content validation of the guide was completed by four subject-matter experts, including two faculty members in public health and two evaluation specialists. Each expert reviewed the guide for clarity, cultural appropriateness, alignment with study objectives, and coverage of key domains. Their recommendations were incorporated into iterative revisions.

For face validation, the guide was pilot-tested in one focus group and two individual interviews with students who met the study’s eligibility criteria but were not included in the final dataset, to avoid contamination of analytic findings. Face validation assessed question clarity, comprehension, flow, and the guide’s ability to elicit the intended constructs. Minor wording adjustments were made to improve readability and conversational flow.

### Survey questionnaire

#### Step 1: Preliminary Item Writing

The survey instrument was developed by a team of three researchers, all trained in quantitative survey design. The team included one PhD-level principal investigator and two master’s-trained research analysts. Survey items were drafted based on existing validated instruments, literature on college well-being, and programmatic priorities. Items were revised through five iterative rounds of review until the full research team reached consensus on content and structure.

#### Step 2: Content Validity

Content validity was assessed by five independent reviewers, including faculty experts in behavioral health, a psychometrics specialist, and two evaluation specialists. Reviewers were provided with the full survey, construct definitions, and domain specifications. Each item was rated for relevance, clarity, and alignment using a 4-point content validity scale (1 = not relevant, 4 = highly relevant). Items with a content validity index (CVI) below 0.80 were revised or replaced.

#### Step 3: Face Validity

Face validity was established through a pilot test with 12 students representing diverse academic years and demographic backgrounds. Participants evaluated readability, item comprehension, survey flow, burden, and perceived sensitivity of questions. Feedback led to minor adjustments, including simplifying wording, reducing redundancy, and improving skip logic.

### Data analysis

#### Survey

Statistical analysis was used to evaluate the relationship between vaccination status (or the desire to receive a vaccine) and relevant demographic characteristics, including gender, age, country, etc. Normally distributed continuous data were summarized by means and standard deviation, while skewed continuous data were summarized by median and interquartile range. Categorical data were summarized by frequency and percentage. All statistical analyses were done at the significance level (alpha) = 0.05, two-sided. P-values less than the alpha value were considered to be statistically significant; otherwise, they were considered not statistically significant. All statistical assumptions were confirmed using appropriate tests.. To conduct these analyses, we utilized the Statistical Analysis System software (SAS 9.4 Institute, Cary, NC) for data cleaning and manipulation, ensuring robust and accurate results. SAS 9.4 and R statistical software 4.11.

#### Focus groups and interviews

Audio-visual recordings of focus group and interview discussions were transcribed manually through approved human transcribers. Two researchers (both with a Ph.D. degree and more than 10 years of experience in qualitative research) have used inductive coding techniques to analyze data. They followed a constant comparative approach to create and condense codes as required. The coding was done manually in Dedoose Version 9.0, and with each new code identified from additional focus groups, the codebook was expanded. The previous coding was reviewed and re-coded based on previously published work. The analysis of focus group discussions reached saturation when no new codes were identified, and themes were repeated. The primary researcher developed the codebook, which was refined through discussion with two senior researchers, and everyone agreed upon the final codebook.

## RESULTS

### Demographics

Demographics and other participant characteristics relevant to the scope of the study are provided in Table 2. More number of participants responded in pre-survey (n=124) than post-survey (n=89).

**Table 2.**
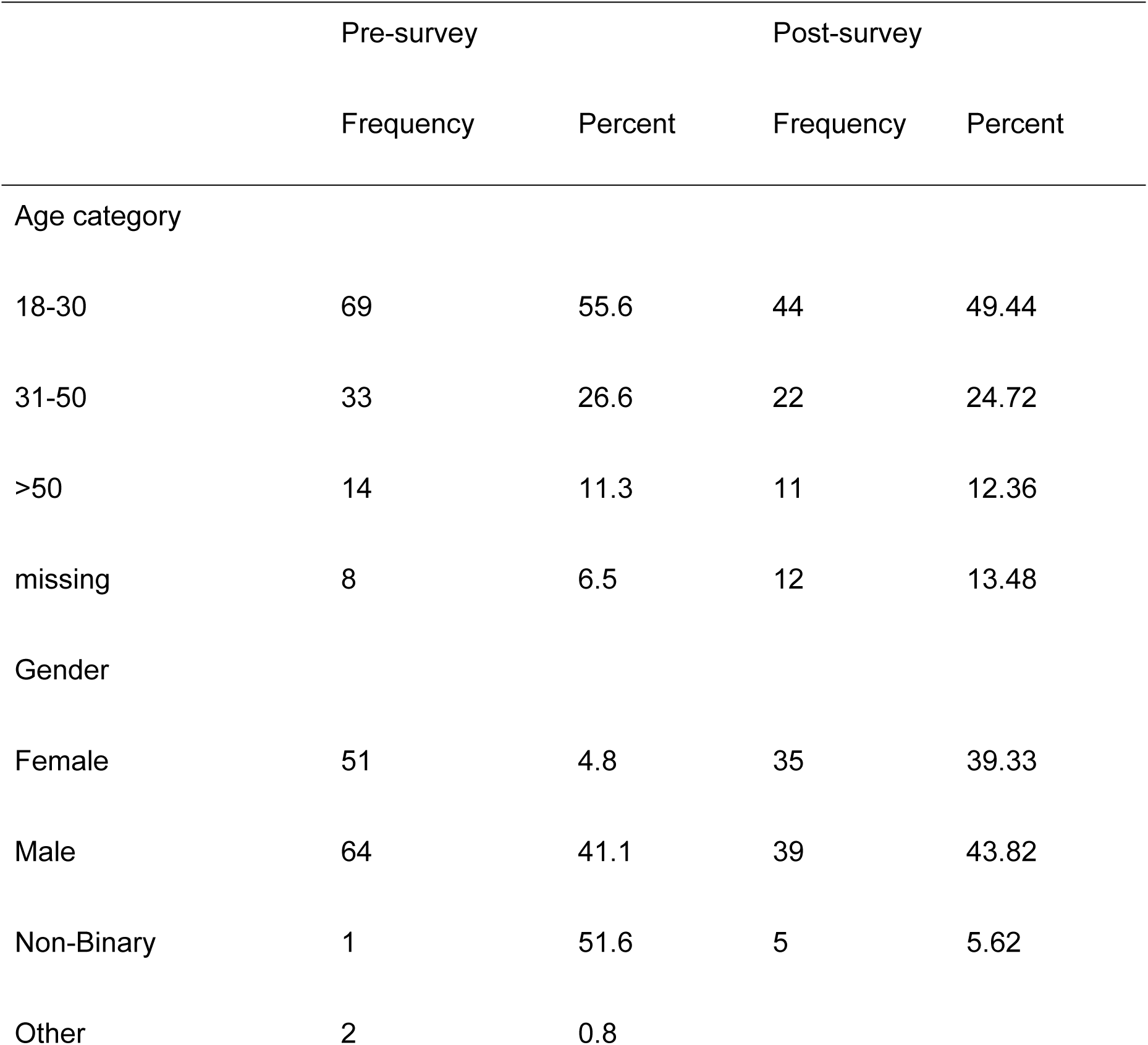
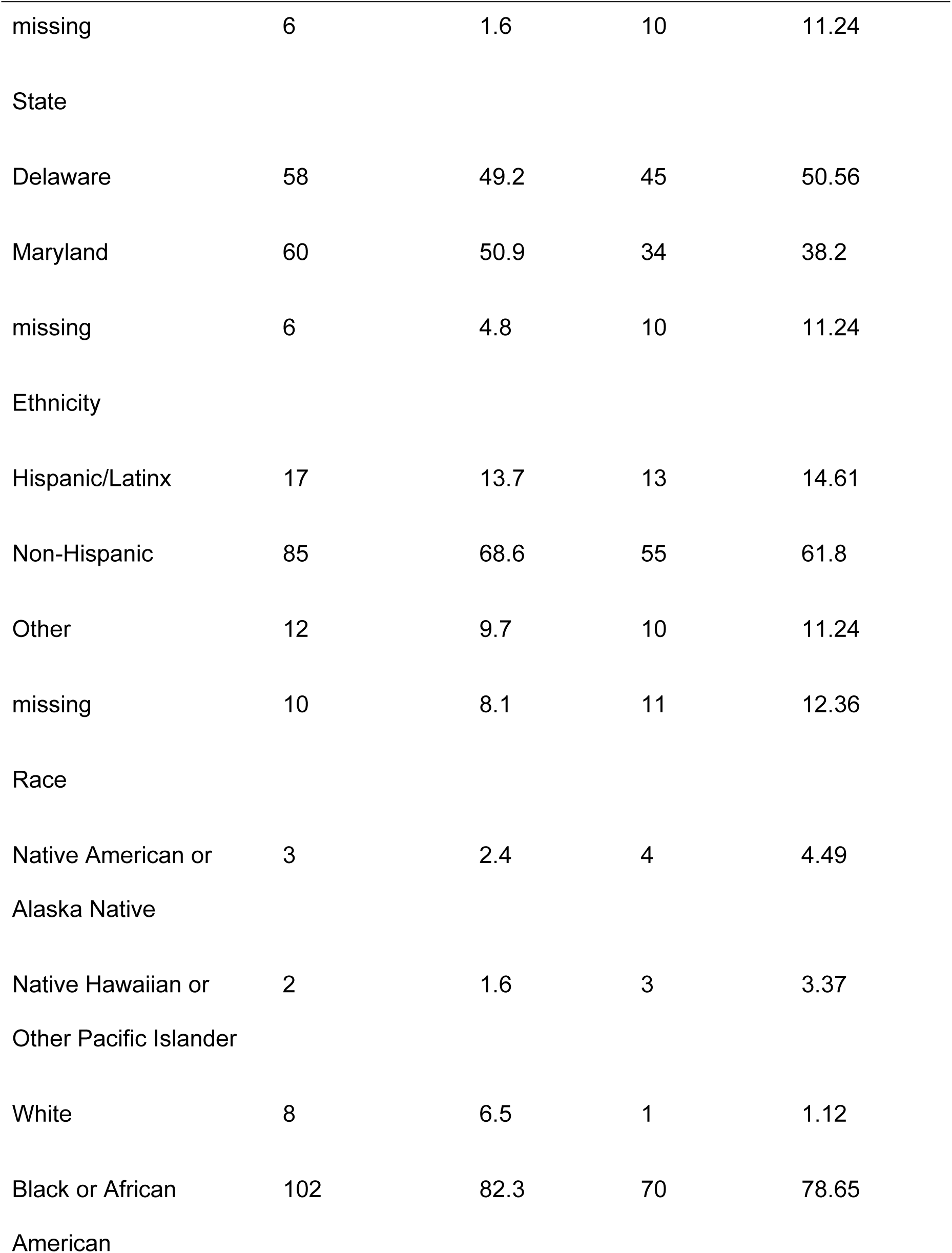
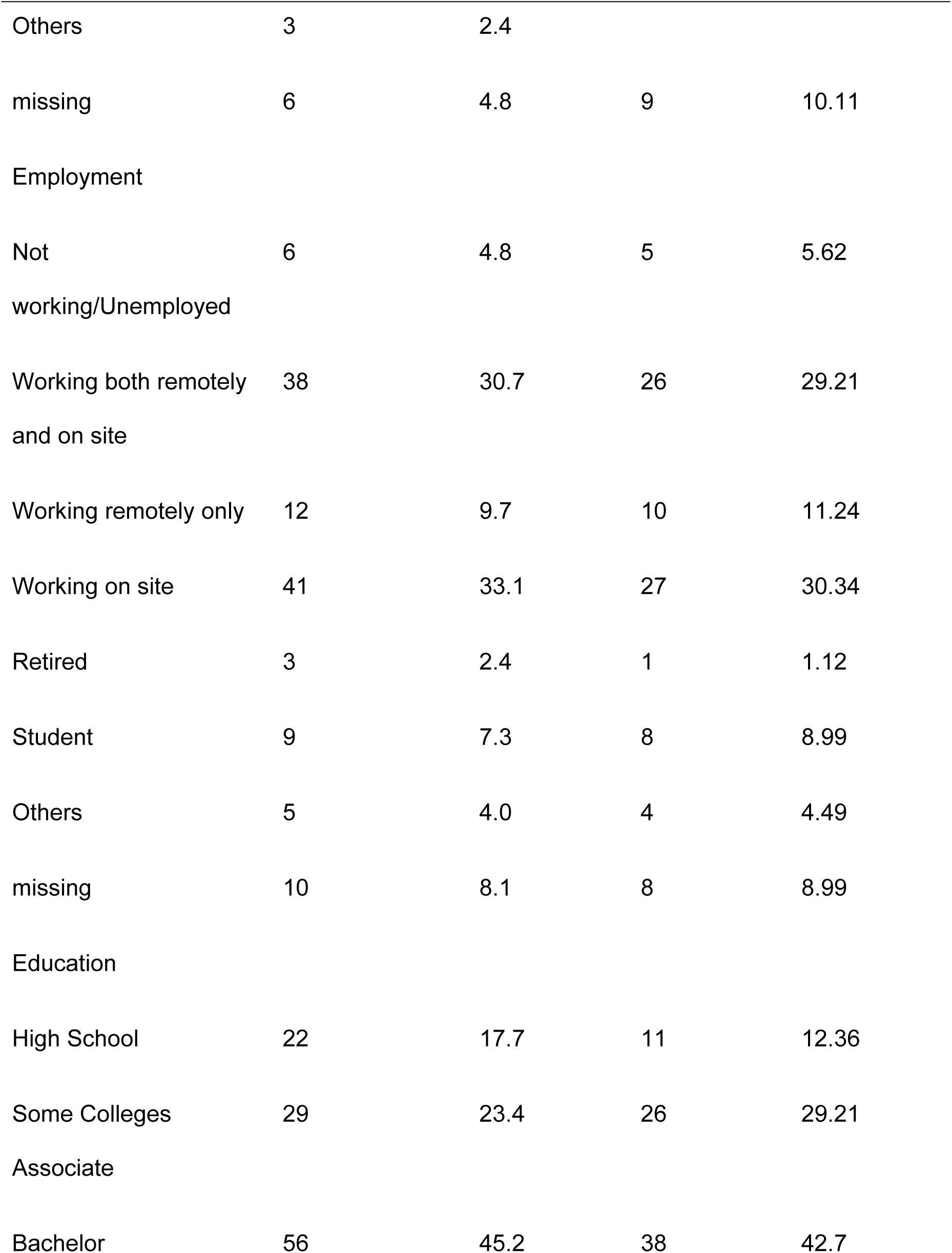
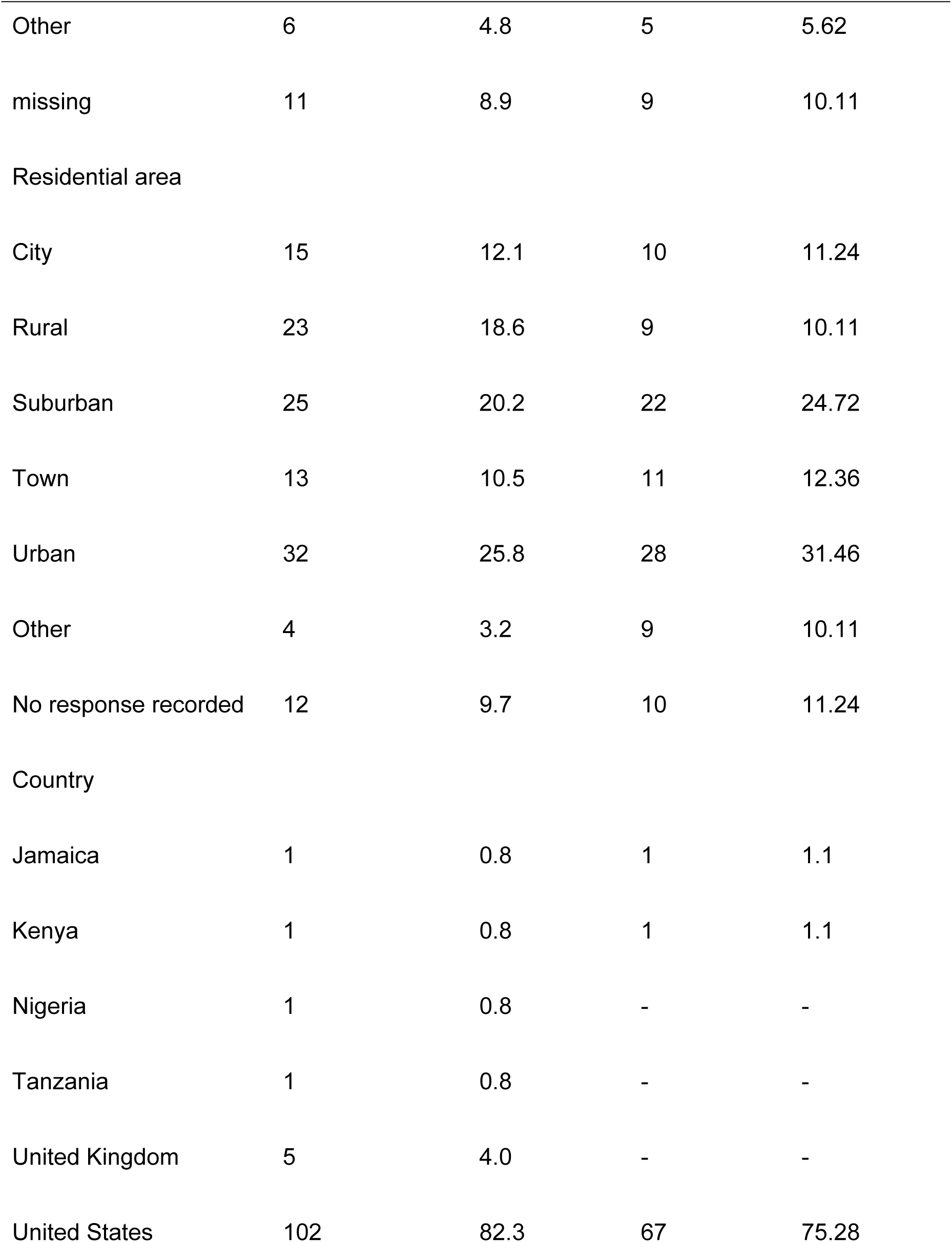
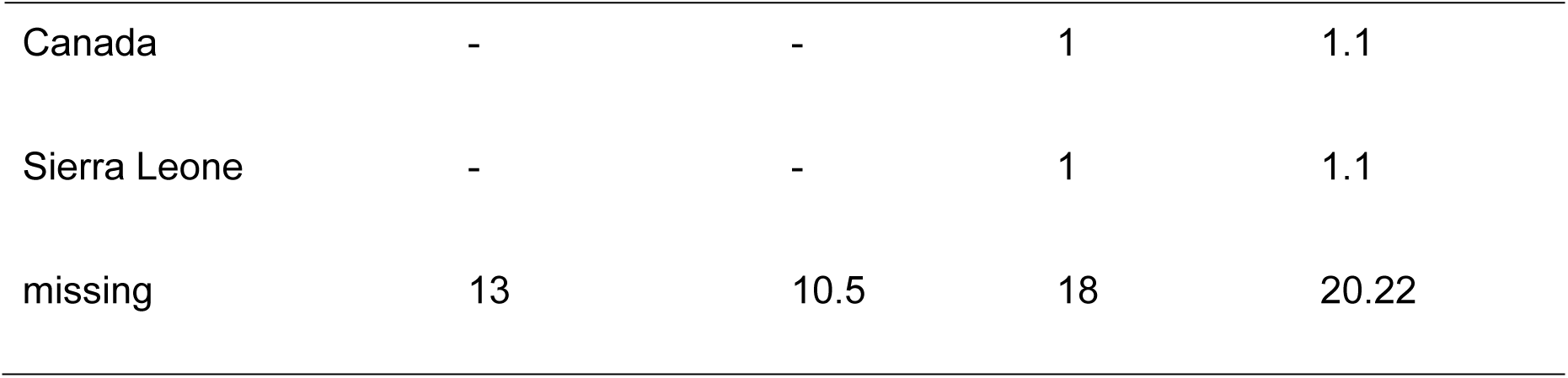
Demographics of the study population pre-survey and post-survey.

Table 3 highlights the educational intervention’s significant impact on participants’ vaccination perceptions. Barriers to vaccination saw a dramatic decline post-intervention (336 pre vs. 23 post), particularly in themes like “Lack of authentic information” and “Behavioral barriers,” reflecting improved clarity and reduced hesitancy. Facilitators for vaccination and perception-related mentions also decreased post-intervention (457 to 45 and 628 to 45, respectively), indicating that participants gained confidence and resolved prior uncertainties. Trust, however, increased substantially (368 pre vs. 170 post), with “Personal health care” (58 to 168) emerging as a key driver, showcasing growing reliance on healthcare providers for guidance. Minimal change in “Willingness of frequency” (2 pre vs. 1 post) suggests that participants’ preferences regarding vaccination schedules remained stable. Overall, the table demonstrates the intervention’s effectiveness in addressing barriers, enhancing trust, and fostering positive perceptions of vaccination.

**Table 3.** Frequency of themes pre-intervention and post-intervention.

|  | Pre-intervention | Post-intervention |
| --- | --- | --- |
| Barriers to vaccination | 336 | 23 |
| Age | 11 | 0 |
| External (other people) experience/information | 26 | 1 |
| Health factor | 65 | 12 |
| Behavioral barrier | 30 | 3 |
| Lack of authentic information | 193 | 7 |
| Vaccination unavailability | 11 | 0 |
| Facilitators for vaccination | 457 | 45 |
| Ease of availability | 8 | 0 |
| Family/ friends | 71 | 5 |
| First-hand experience | 46 | 7 |
| Information on vaccination | 97 | 16 |
| Others safety | 103 | 4 |
| Psychological factors | 75 | 13 |
| Information sources | 57 | 0 |
| Perception | 628 | 45 |
| Encouraged | 14 | 9 |
| Feel safe | 73 | 3 |
| Motivated | 12 | 3 |
| Perception - others | 87 | 0 |
| Perception-children | 72 | 3 |
| Perception-information | 220 | 18 |
| Perception-mandatory vaccination | 62 | 5 |
| Negative perception of mandatory vaccination | 88 | 4 |
| Trust | 368 | 170 |
| Personal health care | 58 | 168 |
| Social media - lack of credibility | 30 | 0 |
| Trust - internet | 22 | 0 |
| Trust - public health agencies | 126 | 0 |
| Trust - science | 124 | 2 |
| Trust- social media | 8 | 0 |
| Willingness of frequency | 2 | 1 |
| As per requirement/suggested | 0 | 1 |
| Not very often | 2 | 0 |
| Totals | 1791 | 284 |

### Pre-intervention

The relationship between vaccination status or desire to receive a vaccine and relevant demographics was explored before and after the implementation of education and awareness implementation. A univariable binomial logistic regression was used to test and evaluate the relationship between vaccination status and relevant demographics. Only children’s vaccination status (whether children received a vaccination or not) and education were significantly associated with the vaccination status of the individuals considered (*p* = 0.0001 for children’s vaccination; *p* = 0.0081 for education). Because of the low event for education, the odds ratio could not be generated. The odds ratio for children vaccination was 0.072, indicating that the chances of individuals who received vaccines and whose children did not receive vaccines are reduced by 92.8 % compared to those who received and whose children received vaccines (*p* = 0001). Although a multivariable binomial regression is required to confirm this association, however the small sample size (low power) coupled with the fact that most of the demographics were not significant in the univariable binomial logistic regression makes this unnecessary.

### Barriers to vaccination

This study focused on perceived inadequacies in available information about vaccines. The recurrent theme across the participant responses was a significant concern about the lack of authentic and comprehensive information regarding vaccinations, which hinders their willingness to get vaccinated. Participants frequently expressed a need for more detailed information about vaccine safety, side effects, long-term effects on health, and the overall efficacy of vaccines in preventing diseases. For instance, one participant stated, *”I want to know what the vaccine is doing to my body in the long term.”* Participants articulated a distrust in the information disseminated through mass media and public health communications, often labeling it as incomplete or misleading. Many indicated that their hesitation to receive vaccines was amplified by their perception that the information provided was not tailored to their personal health needs or did not adequately address their fears about potential adverse effects. A participant noted, *”The information I get from [the media] is not enough for people to get vaccination.”* There was also a notable concern regarding the transparency and reliability of the sources providing vaccine-related information, with many participants expressing a preference for more direct communication from healthcare providers. The lack of detailed and clear information has led to a reliance on personal anecdotes and external experiences, which often skew towards highlighting negative outcomes, further fostering vaccine hesitancy. A participant summed up the sentiment: *”I feel like they don’t give me enough information. I need more information to be sure.”* Additionally, some participants mentioned myths and misconceptions about vaccines, such as fears about infertility and microchipping, which are propagated by insufficiently addressed rumors and lack of authoritative denials or clarifications. *”Some myths have been advanced that maybe you get vaccinated against COVID-19 you may be sterile,”* one respondent commented, illustrating the impact of such myths on vaccination decisions.

### Facilitators for vaccine uptake

This study identifies key facilitators that encourage individuals to seek and obtain vaccinations, with a primary focus on the role of information sources and trusted networks. Participants highlighted a diverse range of information sources, including internet searches, government resources, and social media platforms, as pivotal in their decision-making processes regarding vaccinations. For instance, one participant emphasized the importance of using *”government agencies”* and another trusted *”health provider apps”* as reliable sources. Additionally, personal networks play a critical role in shaping vaccination decisions. Many respondents reported that family, friends, and healthcare providers are significant influencers. One participant mentioned *”the people you trust,”* including classmates and extended family, as crucial in their choice to get vaccinated. Another highlighted the impact of first-hand experiences shared within their social circle, stating, *”my dad’s a doctor,”* underscoring the trust placed in close personal connections. Moreover, participants noted the dual influence of the internet, recognizing its potential for both misinformation and valuable health information. While some expressed skepticism about the credibility of online information, others felt confident in combating misinformation through selective trust in reputable sites and health apps.

### Perception around vaccination

The analyzed data focuses on individual perceptions regarding vaccination, highlighting a complex spectrum of attitudes that range from strong support to significant skepticism. These perceptions are influenced by various factors including personal experiences, the quality of information received, trust in scientific research, and concerns over potential side effects or mandatory vaccination policies. The theme reflects a dynamic interplay among access to reliable information, personal autonomy, community health responsibilities, and trust in health authorities.

#### Positive Perceptions

Participants who viewed vaccination positively often cited reliable sources of information and beneficial personal experiences with vaccines. There is a clear recognition of the role vaccines play in preventing serious illnesses and in safeguarding community health. Some individuals expressed a strong sense of duty towards community safety, advocating for mandatory vaccination in settings like schools or for international travel. Trust in the scientific process and the effectiveness of established vaccines also contributed to favorable perceptions, underscoring a reliance on scientific authority and proven health measures.

#### Negative Perceptions

Conversely, skepticism about vaccine efficacy and safety was frequently mentioned, particularly concerning the rapid development and approval of new vaccines. Many expressed apprehensions about potential side effects and a perceived lack of transparent, authentic information, which acted as significant barriers to vaccine acceptance. The concept of mandatory vaccination was particularly contentious, with numerous respondents emphasizing personal freedom and ethical concerns over compulsory health measures.

Additionally, mistrust in health information, especially from unofficial sources and social media, led to reluctance or outright refusal to engage with vaccination programs.

### Trust

The theme of “Trust” in public health agencies and science in relation to vaccination is prevalent, with respondents expressing varying levels of confidence. This theme captures individual sentiments towards the reliability and expertise of public health agencies and the scientific community in handling vaccines. It encompasses a broad spectrum of trust, from complete confidence due to the professional training and informational accuracy provided by these bodies, to cautious skepticism influenced by past experiences and the rapid development of new vaccines.

A significant majority of participants expressed strong trust in public health agencies, emphasizing that the training and expertise of these organizations make them reliable sources for vaccination guidance and public health safety. Many participants pointed out that the agencies’ responsibilities to public health and their role in controlling diseases through vaccination justify their trustworthiness. For instance, one respondent stated, *”Yeah, I think the public agencies should be trusted since they have a good knowledge on the vaccination, so I think they should be trusted.”* However, some reservations were noted, particularly regarding the rapid approval and recommendation of new vaccines, which stirred some concerns about transparency and the influence of external agendas. A participant highlighted, *”I think they’re effective because they will have been tested over some period of time and they will have determined if it works or not.”*

Trust in science was predominantly positive, with many respondents acknowledging that their confidence in getting vaccinated was bolstered by their trust in scientific research and the effectiveness of vaccines proven through scientific methods. The ability of vaccines to prevent diseases and protect public health was a recurring rationale for this trust. For example, a participant mentioned, *”I think, yes, because vaccine protects you from contracting a disease before it makes you sick, yeah.”* Nonetheless, a few participants expressed a more measured trust, contingent on the availability of transparent and comprehensive information about vaccine development and side effects, which they felt were sometimes lacking. Reflecting this sentiment, another respondent remarked, *”Well, yes, I think that vaccinations can be effective because it creates immune response against such diseases.”*

### Willingness of frequency

The theme “Willingness of Frequency” explores participants’ openness to the frequency of receiving vaccinations, particularly boosters, as a reflection of their trust in vaccine efficacy and their personal health philosophies. This theme illustrates the varied attitudes towards ongoing vaccination schedules, weighing personal health maintenance against concerns for potential over-vaccination. Participants who displayed a positive willingness towards frequency typically expressed a readiness to receive vaccines regularly, with several even specifying intervals such as twice a year or annually. A common sentiment among this group was the desire to stay protected and safe, with one participant stating, *”Often. Regularly, as long as it keeps me safe and protected.”* The assurance of safety and the acknowledgment of vaccines’ role in maintaining health were pivotal in their willingness to receive vaccines more frequently. Some were agreeable to following booster schedules, as indicated by one participant: *”As often as I need it because I am boosted.”*

Conversely, some participants were hesitant or outright resistant to frequent vaccinations. Their reluctance often stemmed from a distrust in the necessity of multiple booster shots, needle aversion, or a belief in the longevity of vaccine protection. Skepticism about the motivations behind booster recommendations was also evident, as some felt the push for boosters was excessive and profit-driven. The notion of once-and-done vaccinations resonated with this group, as echoed in one comment: *”I wouldn’t say every year. [laughter] Every few years maybe, but not all the time.”*

### Post-intervention

Similarly, for the post-survey part, a univariable binomial logistic regression was used to test and evaluate the relationship between vaccination status and relevant demographics, as shown in Table 3. Only the desire to have children (whether a participant has children or not, yes/no) and Education were significantly associated with the vaccination status of the individuals considered (*p* = 0.0001 for children vaccination; *p* = 0.0302 for education). Because of the low event for education, the odds ratio could not be generated.

### Barriers to vaccination

The theme “Barriers to Vaccination Post-Education Intervention” explores the multifaceted reasons why individuals remain hesitant or refuse vaccinations even after being provided with educational information about their benefits. These barriers are not merely a lack of information but are deeply intertwined with personal health beliefs, experiences, psychological factors, and perceived authenticity and efficacy of the vaccines. Individuals express concerns that vaccinations are not a *”one size fits all”* solution and emphasize the importance of personal health needs and the individual’s right to choose based on a thorough understanding of potential risks and benefits. Experiences of adverse reactions, either first-hand or within one’s community, significantly contribute to this hesitancy, further compounded by mixed messages regarding the necessity and frequency of booster shots. This suggests that while education can inform, it cannot fully overcome the lived experiences and personal beliefs that shape one’s attitude toward vaccination. Conversely, a notable portion of the group expresses skepticism or outright rejection of further vaccinations, especially booster shots. This stance is fueled by concerns about the experimental nature of some vaccines, the lack of long-term data, personal or observed adverse reactions, and a strong belief in autonomy over one’s health choices. For example, one participant noted, *“I’m not understanding, which makes me feel like, ‘Okay, well, so what they’re injecting us is not lasting long, and that’s why you have to continue to keep getting the boosters.’”* Concerns were raised about the necessity of boosters, suggesting doubt about the longevity of vaccine protection and the robustness of the vaccination process itself. One participant mentioned how their family member, who got vaccinated *“…and they were unable to walk for an extended time after that.”*

### Facilitators for vaccine uptake

The theme “Facilitators for Vaccination Post-Education Intervention” highlights the positive influence of educational resources, such as fact sheets and informational videos, on individuals’ attitudes toward vaccination. These resources served as key facilitators by providing comprehensive, credible, and accessible information that helped to demystify vaccines, elucidate their importance across various age groups, and clarify their role in public health. Participants underscored the value of clear, concise, and factual content in empowering them to make informed decisions. The intervention successfully engaged diverse individuals, addressed their concerns, and often positively changed their perspectives toward vaccinations. A segment of the participants maintains a positive view of vaccinations, often influenced by personal health conditions (e.g., diabetes) and recommendations from trusted community figures like religious leaders. One participant noted, *”Because the preacher said he has diabetes, so he’s faithful and taking his insulin shot. And he encourages his congregation… to consider the vaccinations for health reasons.”* Participants reported an increased understanding of vaccines due to the educational materials provided. *”The fact sheet and the video… made me realize the importance of getting vaccinated.”* The clarity and comprehensiveness of the information helped dispel myths and encouraged open discussion, leading to more positive attitudes towards vaccination. The credibility of the information presented was frequently mentioned as a facilitator for considering vaccination. Participants expressed appreciation for the presentation of facts by professionals and trusted sources, which contributed to the credibility and acceptance of the message. *”The video… had people of different ages and professions giving their experience on vaccination.”* The diversity in perspectives presented in the video, including age, profession, and personal experiences, resonated with the audience and allowed for broader identification and relatability. *”It is very diverse… You have different representation in the video… It is for everyone, and it is for everyone to live a better life.”* The easy accessibility of the educational resources and the simplicity of the information presented were crucial in encouraging individuals to consider vaccination. Participants valued being able to easily access detailed information about different vaccines and their specific age recommendations.

### Perception around vaccination

The theme “Perception Post-Education Intervention” encapsulates participants’ viewpoints and sentiments following exposure to educational content on vaccinations. The post-intervention perceptions display a spectrum ranging from highly encouraged and enlightened to cautious and skeptical. This diverse array of personal conclusions and attitudes towards vaccines reflects the multifaceted impact of the intervention. While some individuals became motivated to act based on newfound knowledge, others retained reservations, emphasizing the need for personal choice and further inquiry. Many participants experienced a positive shift in perception, reporting increased interest and motivation to seek vaccinations due to the informative nature of the intervention. The educational content was praised for its inclusivity and representation, making the information relatable and trustworthy. *”The video is contained with a--it represents black people… so the black people, indigenous, and people of color would know that they are also entitled to get vaccines.”* Participants felt empowered to make enlightened decisions regarding their own health and that of their families. The intervention’s success was attributed to its ability to clarify the relevance of specific vaccines for different age groups and personal health conditions. *”I know the kind of vaccine that I should get for my age and for my kids’ age and I think that is just one very, very great advancement for me in knowledge.”* Despite the overall positive feedback, there was a strong sentiment for the need to maintain personal choice in health decisions. Some participants were wary of perceived coercion, particularly in the context of mandates, and valued the freedom to choose based on individual beliefs and circumstances. *”Everyone should be able to make their own choice as far as being vaccinated.”* A segment of the audience remained cautious and skeptical, indicating that while the intervention was informative, it was not entirely persuasive. Some expressed concerns about the vaccines’ efficacy and the potential for adverse reactions. *”Vaccinated people still get sick and they still spread to unvaccinated people, so I don’t really think it prevents it.”*

### Trust

The theme “Trust Post-Education Intervention” refers to the establishment and reinforcement of credibility and confidence among participants in the information provided about vaccines. This theme emerged after the participants were exposed to an educational intervention comprising videos and fact sheets that aimed to broaden their understanding of various vaccines beyond just COVID-19. Participants evaluated the content’s reliability, expressed how it resonated with their personal experiences, and considered its applicability to their lives. Participants expressed a heightened sense of trust due to the credible information presented in the materials. The videos and fact sheets were particularly effective when they aligned with personal experiences or when they saw their perspectives reflected in the content. *”And both the video and the fact sheet made me feel safer about vaccines, and I definitely feel more knowledgeable.”* The educational content successfully expanded participants’ awareness of vaccinations beyond COVID-19, introducing them to a range of other important vaccines and demonstrating the lifelong nature of vaccination. *”And it’s also very educative since, at the moment, many infectious diseases are appearing as a result of vaccinations, and some are still coming up.”* Several participants suggested ways to improve the dissemination of vaccine information, such as incorporating children in the educational materials, using larger posters for public awareness, and providing additional context for diseases and their prevention through vaccination. *”Maybe a little information on the disease will be helpful.”* Diverse perspectives shared in the video component helped validate personal feelings and encouraged informed decision-making based on the testimony of others from various walks of life. *”Everyone has given an opportunity to give their own opinions… Then they were educated on the importance of the vaccines.”* Exposure to information through the intervention swayed some participants towards considering more vaccinations for themselves and their families, indicating an increased level of trust in the vaccines’ purposes and safety. *”I would try to want to get vaccinated more… It is an encouragement.”*

### Willingness of frequency

The theme “Willingness to Vaccinate” reflects participants’ conditional openness to receiving vaccines based on factors such as the purpose of the vaccine, the extent of research conducted, and the availability of essential information. While some participants expressed hesitancy about being early adopters, their willingness to adopt could increase if the vaccine is necessary for survival or supported by thorough research and credible information. *“I’m not going to be first in line to get it. If I get it, I need to do my research. I need more information first.”*

## DISCUSSION

The study explored participants’ perceptions regarding vaccination, identified strategies to increase vaccine confidence through community engagement, and assessed changes in perceptions post-intervention. This educational intervention, focusing on increasing vaccine uptake among older adults, demonstrates several significant outcomes that highlight the complexity of vaccine acceptance and the effectiveness of tailored educational strategies. The results indicate that participants gained enhanced understanding and appreciation of vaccine-related knowledge through the comprehensive information presented in the videos and fact sheets.

### Pre-intervention findings

The pre-intervention results underscore the importance of children’s vaccination status and educational attainment in influencing individual vaccination behaviors. The significant association between vaccination status and these variables aligns with existing research emphasizing the role of family and educational context in shaping health decisions. For example, studies have demonstrated that parents’ vaccination decisions are closely tied to their trust in vaccines for their children, reflecting broader societal and cultural influences (Dubé et al., 2015). The barriers identified, such as lack of comprehensive information and distrust in sources, are well-documented contributors to vaccine hesitancy (Larson et al., 2016). Participants’ concerns regarding long-term effects and transparency are consistent with other studies highlighting the need for tailored, clear communication to address vaccine safety concerns (MacDonald, 2015). Myths and misconceptions, such as fears of infertility and microchipping, illustrate the pervasive impact of misinformation, which has been exacerbated during the COVID-19 pandemic (Loomba et al., 2021). The study revealed that trusted networks, including healthcare providers, family, and community leaders, were pivotal in shaping vaccination decisions. This finding aligns with previous research emphasizing the importance of leveraging trusted messengers in public health campaigns to enhance vaccine uptake (Quinn et al., 2017). Additionally, participants’ acknowledgment of the internet and health apps as sources of both information and misinformation underscores the dual role of digital platforms. This highlights the necessity for public health agencies to actively counter misinformation while promoting credible online resources (Chou et al., 2021).

### Post-intervention changes

The educational intervention demonstrated a positive impact on participants’ perceptions and willingness to vaccinate. The use of videos and fact sheets to demystify vaccines and address specific concerns reflects best practices in health communication (Freimuth et al., 2017). Notably, participants praised the inclusivity of the materials, which featured diverse representation and relatable narratives. This inclusivity likely enhanced the intervention’s effectiveness by fostering trust and relatability, consistent with findings that culturally tailored interventions are more successful in reducing hesitancy (Habib et al., 2023). Despite these positive changes, some participants remained skeptical, citing concerns about the necessity and frequency of boosters and mistrust in rapid vaccine development. This aligns with prior studies showing that while education can reduce hesitancy, deeply ingrained beliefs and experiences often require sustained engagement to shift (Yaqub et al., 2014). Trust emerged as a central determinant of vaccination attitudes, both pre- and post-intervention.

Participants’ mixed levels of trust in public health agencies and scientific processes are reflective of broader societal trends, where confidence in institutions varies widely (Larson et al., 2016). While many participants expressed increased trust following the intervention, lingering doubts underscore the need for transparency and consistent messaging to rebuild public confidence (Betsch et al., 2015). Highlighting the long-term benefits and safety profiles of vaccines, along with addressing perceived coercion, may further enhance trust and acceptance.

Participants not only increased their knowledge but also felt reassured about the safety and longstanding use of vaccines, echoing previous findings that trusted information can reduce vaccine hesitancy (Brewer, 2021; Habersaat & Jackson, 2020; Joslyn et al., 2023; Rani et al., 2022; Reiss, 2022). However, the intervention also underscores the individuality of vaccine acceptance. While many participants recognized the critical role of vaccines in disease prevention and health maintenance, personal beliefs and preferences played a crucial role. A subset of participants maintained that vaccination should remain a personal choice, supporting literature that suggests personal health freedoms are a significant factor in vaccine decision-making (Agranov et al., 2021; Canning et al., 2022; Slotte et al., 2022; Sprengholz et al., 2021). This highlights the necessity of respecting individual autonomy while designing public health campaigns.

Notably, our intervention addressed potential disparities in vaccine acceptance and knowledge by including diverse representation within the educational material. This inclusivity may have strengthened the relevance and impact of the information across different racial and ethnic groups, which is often overlooked in public health initiatives (AuYoung et al., 2023; CDC, 2022; Koku et al., 2024; Richard-Eaglin & McFarland, 2022; Upshaw et al., 2024). The positive feedback on representation suggests that culturally sensitive approaches are crucial in enhancing the effectiveness of health interventions.

Despite these advances, the intervention illuminated the limits of information dissemination alone. Some participants indicated that the information, while valuable, would not alter their existing beliefs or vaccination intentions. This aligns with findings from previous studies, which noted that deeply held beliefs and past experiences significantly shape health behaviors, potentially outweighing new information (Brennan-Ing et al., 2023; Brewer, 2021; Chu & Liu, 2021; Shahrabani & Benzion, 2012; Xu et al., 2021). Moreover, the intervention highlighted the diverse responses to vaccine information, influenced by past personal experiences with vaccines and the perceived experimental nature of some vaccines. Such individual differences suggest the need for a more personalized approach to public health messaging that addresses the specific concerns and backgrounds of different demographic groups.

### Implications for Public Health

The findings suggest several actionable strategies to reduce vaccine hesitancy:

- Community Engagement: Leveraging trusted community leaders and networks to disseminate vaccine information can enhance credibility and acceptance.
- Culturally Tailored Interventions: Developing materials that reflect the diversity of target populations increases relatability and effectiveness.
- Enhanced Communication: Addressing safety concerns transparently, providing long-term efficacy data, and debunking myths with evidence-based information are crucial.
- Sustained Efforts: While one-time interventions are beneficial, ongoing education and engagement are necessary to address deep-seated skepticism and misinformation.

## CONCLUSION

The findings from this study advocate for the development of educational materials that not only convey crucial health information but also engage with the audience’s values and beliefs. Accessibility of information was notably enhanced in this intervention, making it easier for older adults to understand and engage with the content, which is essential for informed decision-making. As participants valued the ability to use the information as a tool to educate others, this suggests that peer influence and community engagement might be effective strategies for further enhancing vaccine uptake. This study contributes to the broader discourse on public health strategies for increasing vaccine uptake, emphasizing the importance of tailored, respectful, and inclusive educational approaches to address the multifaceted nature of vaccine hesitancy and acceptance. Further research should explore the long-term impact of such interventions and the potential for integrating these strategies into larger populations to maximize public health benefits.

### Limitations

The current study is significantly underpowered (below 80%), which makes it difficult to detect meaningful associations between vaccination status and relevant variables and demographics. It is crucial to conduct a power analysis before any observational studies to determine the appropriate number of subjects or participants needed to achieve the required statistical significance or effect size. Currently, this study’s low sample size limits its power. Despite this limitation, the study has notable strengths, including its emphasis on community engagement and its dual evaluation of perceptions both before and after the intervention.

However, the small sample size restricts generalizability, and the reliance on self-reported data may introduce bias. Future studies should aim to include larger samples.

## Data Availability

The study includes mixed-methods data; quantitative results are fully reported in the manuscript, while qualitative data are not publicly available due to ethical and confidentiality protections.

## Acknowledgments

The authors acknowledge the contributions of students and staff at Delaware State University and UMES Extension to data collection efforts.

Funding for this study was provided through the Extension Collaborative on Immunization, Teaching & Engagement (EXCITE)-1890 Universities Foundation project.The funding agency had no role in the study design, data collection, analysis, interpretation, or preparation of this manuscript.

## Disclaimer

The authors are solely responsible for the study design, data collection, analysis, interpretation of findings, and conclusions presented in this article. The views expressed do not necessarily reflect those of any affiliated institutions, funding organizations, or government agencies.

